# Immunogenicity and reactogenicity of a heterologous COVID-19 prime-boost vaccination compared with homologous vaccine regimens

**DOI:** 10.1101/2021.06.13.21258859

**Authors:** Tina Schmidt, Verena Klemis, David Schub, Janine Mihm, Franziska Hielscher, Stefanie Marx, Amina Abu-Omar, Sophie Schneitler, Sören L. Becker, Barbara C. Gärtner, Urban Sester, Martina Sester

**Affiliations:** Department of Transplant and Infection Immunology, Saarland University, 66421 Homburg, Germany; Department of Internal Medicine IV, Saarland University, 66421 Homburg, Germany; Institute of Medical Microbiology and Hygiene, Saarland University, 66421 Homburg, Germany

## Abstract

Heterologous priming with the ChAdOx1-nCoV-19 vector-vaccine followed by boosting with an mRNA-vaccine is currently recommended in Germany, although data on immunogenicity and reactogenicity are not available. Here we show that the heterologous regimen induced spike-specific IgG, neutralizing antibodies, and spike-specific CD4 T-cells, which were significantly more pronounced than after homologous vector boost, and higher or comparable in magnitude to the homologous mRNA regimens. Moreover, spike-specific CD8 T-cell levels after heterologous vaccination were significantly higher than after both homologous regimens. Cytokine expression profiling showed a predominance of polyfunctional T-cells expressing IFNγ, TNFα and IL-2 with subtle differences between regimens. Both recipients of the homologous vector-regimen and the heterologous vector/mRNA-combination were most affected by the priming vector-vaccination, whereas heterologous boosting was well tolerated and comparable to homologous mRNA-boosting. Taken together, heterologous vector-mRNA boosting induces strong humoral and cellular immune responses with acceptable reactogenicity profile. This knowledge will have implications for future vaccine strategies.

## Main Text

Among the currently licensed coronavirus disease (COVID-19) vaccines, the ChAdOx1 nCoV-19 adenovirus-based vector-vaccine (ChAdOx1) and the two mRNA vaccines (BNT162b2 or mRNA-1273) have been most widely applied. Both vaccine types are immunogenic and have shown remarkable efficacy in preventing COVID-19^1-3^. In March 2021, administration of the ChAdOx1 vaccine was temporarily suspended in Germany due to the occurrence of life-threatening cerebral venous thrombosis and thrombocytopenia primarily in younger women^4,5^, a syndrome that was subsequently termed as vaccine-induced immune thrombotic thrombocytopenia (VITT). This resulted in revised recommendations for secondary vaccination of all individuals who had received the first dose of the vaccine. Individuals above the age of 60 years are recommended to complete vaccination with the vector-vaccine, whereas heterologous boosting with an mRNA vaccine was recommended below 60 years, with the option to voluntarily remain on a homologous vector regimen^6^. Comparative analyses of immunogenicity between the licensed vaccine regimens are scarce, and knowledge on immunity and reactogenicity after heterologous vaccination is currently limited. We have previously found that priming with the ChAdOx1 vaccine showed a stronger induction of spike-specific T-cell responses as compared to mRNA-priming. In contrast, antibody responses were more pronounced after mRNA-priming^7^. It is currently unknown how differences among the vaccine types after priming may influence cellular and humoral immunity after secondary vaccination. We therefore prospectively enrolled three groups of individuals to study immunogenicity and reactogenicity of a heterologous vector/mRNA prime-boost regimen in comparison to the standard homologous regimens. A detailed analysis of spike-specific IgG-levels and neutralizing antibody activity was performed. In addition, spike-specific CD4 and CD8 T cells were characterized using flow-cytometry. Adverse events within the first week after the priming and boosting dose were self-reported based on a standardized questionnaire.

A total of 216 immunocompetent individuals primarily including employees were prospectively enrolled at Saarland University Medical Center after secondary vaccination with the licensed vaccines ChAdOx1 nCoV-19, BNT162b2, and mRNA-1273. Among study participants, 97 had received heterologous vaccination with the ChAdOx1-vector and mRNA boost (vector/mRNA), whereas 55 and 64 had received homologous regimens with vector or mRNA vaccines, respectively (vector/vector and mRNA/mRNA, table S1). As per guidelines, the time between primary and secondary vaccination was shorter for mRNA-primed (4.3±1.1 weeks) than for vector-primed individuals with no difference between vector-based heterologous (11.2±1.3 weeks) and homologous regimens (10.8±1.4 weeks). Blood samples were drawn at a median of 14 (range 13-18) days after vaccination. All individuals had no known history of SARS-CoV-2 infection. The groups were matched regarding gender; however, individuals on the homologous vector regimen were slightly older than the two other groups who had similar age (table S1). Leukocyte counts including granulocytes, monocytes and lymphocytes as well as major lymphocyte subpopulations such as CD4- and CD8 T cells, and B cells did not differ between the groups (table S1).

Spike-specific IgG were induced in 215/216 individuals after vaccination. Interestingly, IgG-levels after heterologous vaccination were similar as in the homologous mRNA vaccine group (3602 (IQR 3671) and 4932 (IQR 4189) BAU/ml), whereas IgG-levels after homologous vector vaccination were significantly lower (404 (IQR 510) BAU/ml, p<0.0001, figure 1A). This difference was also observed for neutralizing antibody activity, which was quantified by a surrogate neutralization test. While the majority of individuals in the vector/mRNA and mRNA/mRNA group had 100% inhibitory activity, this was significantly lower in the vector/vector group (figure 1B).

**Figure 1:**
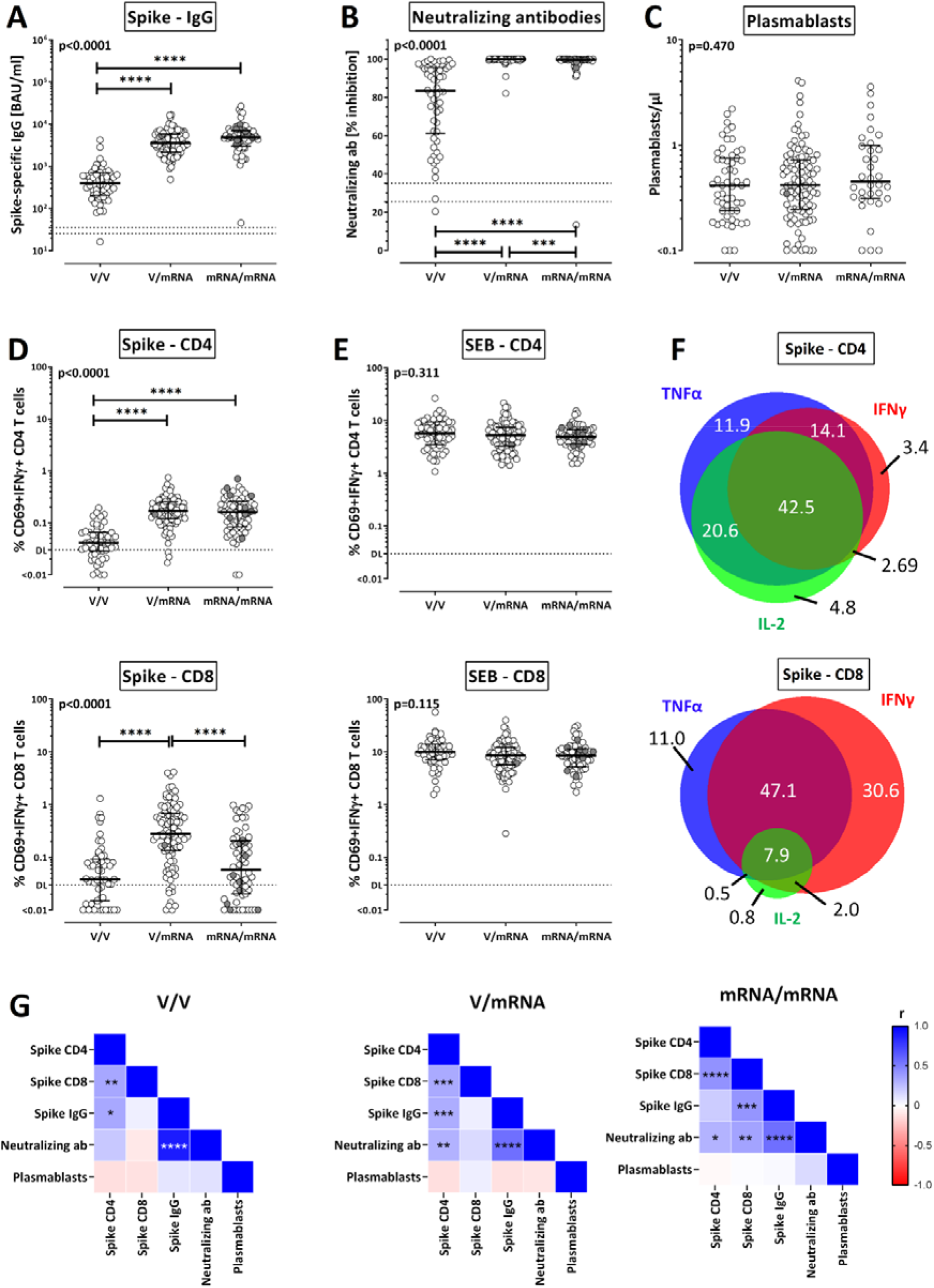
Immune responses towards the SARS-CoV-2 spike protein after vaccination with homologous and heterologous prime-boost regimens. Immune responses were compared between individuals who either received homologous ChAdOx1 nCoV-19 vector vaccination (V/V, n=55), heterologous ChAdOx1 nCoV-19 vector/mRNA vaccination (V/mRNA, n=97) or homologous mRNA vaccination (mRNA/mRNA, n=64). Spike-specific IgG-levels **(A)** and neutralizing antibodies **(B)** were quantified by ELISA and neutralization assay and compared between groups. **(C)** Numbers of plasmablasts were quantified and compared between groups (n=55 for V/V, n=97 for V/mRNA and n=34 for mRNA/mRNA). Percentages of SARS-CoV-2 spike-specific **(D)** and SEB-reactive **(E)** CD4 and CD8 T cells were determined after antigen-specific stimulation of whole blood samples followed by intracellular cytokine analysis using flow cytometry. Reactive cells were identified by co-expression of CD69 and the cytokine interferon (IFN) γ among CD4 or CD8 T cells and subtraction of background reactivity of respective negative control stimulations. **(F)** Cytokine expression profiles of spike-specific CD4 and CD8 T cells in all individuals showing single or combined expression of the cytokines IFNγ, interleukin (IL) 2 and tumor necrosis factor (TNF) α. **(G)** Correlations between spike-specific T-cell levels, antibody responses and numbers of plasmablasts. Bars in (A)-(E) represent medians with interquartile ranges. Individuals who received the mRNA-1273 vaccine are indicated by grey symbols (1 in the heterologous V/mRNA group, 10 in the homologous mRNA/mRNA group). Differences between the groups were calculated using Kruskal-Wallis test with Dunn’s multiple comparisons post test. Correlations in (G) were analyzed according to Spearman. Dotted lines indicate detection limits for IgG in (A) and (B), indicating negative, intermediate and positive levels or levels of inhibition, respectively as per manufacturer’s instructions and detection limits for SARS-CoV-2-specific CD4 T cells in (C) and (D). To allow for robust statistics, analysis in (F) was restricted to samples with at least 30 cytokine-positive T cells (n=181 for CD4 and n=113 for CD8 T cells). SEB, *Staphylococcus aureus enterotoxin* B; *p<0.05, **p<0.01, ***p<0.001, ****p<0.0001.

We have previously shown that induction of primary SARS-CoV-2-specific immunity after natural infection was associated with an expansion of circulating plasmablasts, which correlated with antibody levels^8^. We therefore analyzed whether the different vaccine regimens were associated with differences in the number of plasmablasts, which were identified as CD38-positive cells among IgD^-^CD27^+^ CD19-positive switched-memory B cells (figure S1). The median number of plasmablasts reached 0.42 (IQR 0.49)/µl with no differences between the three regimens (figure 1C). As plasmablast numbers were found to be higher after primary vaccination (median 0.74 (IQR 0.79)/µl)^7^, this may indicate that the boosting dose does not lead to a further increase.

To analyze vaccine-induced T-cell responses, overlapping peptide pools derived from the spike protein were used to stimulate whole blood samples. Spike-specific CD4 and CD8 T cells were identified using flow-cytometry by induction of CD69 and cytokines IFNγ, TNFα and IL-2. The peptide diluent was used as negative control, and polyclonal T-cell reactivity after stimulation with *Staphylococcus aureus* enterotoxin B (SEB) served as positive control for T-cell functionality unaffected by the vaccine regimens. The gating strategy and representative contour plots of cytokine-positive CD4 and CD8 T cells of a 37-year old woman after heterologous vaccination are shown in figure S2. As exemplified for CD69-positive IFNγ-producing T cells, both the heterologous vector/mRNA and the homologous mRNA/mRNA regimen led to a marked induction of spike-specific CD4 T cells with median percentages of 0.17% (IQR 0.13%) and 0.16% (IQR 0.17%), whereas CD4 T-cell levels after homologous vector/vector vaccination were significantly lower (median 0.04% (IQR 0.04%), figure 1D, each p<0.0001). Interestingly, heterologous mRNA boosting in vector-primed individuals induced highest percentages of spike-specific CD8 T cells (0.28% (IQR 0.55%)), which were not only more pronounced than after homologous vector boost (vector/vector: 0.04% (IQR 0.08%)) but also higher than the mRNA/mRNA regimen (0.06% (IQR 0.19)%, p<0.0001, figure 1D). SEB reactive CD4 or CD8 T-cell levels did not differ between the groups (figure 1E). As with IFNγ-producing CD4 and CD8 T cells, similar between-group differences were found for spike-specific CD4 T cells producing TNFα or IL-2, and for spike-specific CD8 T cells producing TNFα (figure S3). As CD8 T cells generally produce less IL-2, differences were less pronounced for IL-2-producing CD8 T cells. Finally, between-group differences were similar if CD4 or CD8 T cells producing any of the three cytokines were considered after Boolean gating (figure S3).

An overview summarizing correlations between spike-specific IgG and their neutralizing activity, spike-specific CD4 and CD8 T cells, and plasmablasts is shown in figure 1G. As expected, IgG-levels showed a significant correlation with neutralizing activity in all three groups. A strong correlation was also found for spike-specific CD4 and CD8 T-cell levels. In line with a role of CD4 T cells in supporting antibody production, CD4 T cells correlated with IgG-levels in both the vector/vector and the vector/mRNA group. In the mRNA/mRNA group, antibody levels and neutralizing activity were found to correlate with CD8 T-cell levels. Whether this reflects a causal relationship or similar induction kinetics is currently unknown.

Apart from quantitative analysis of spike-specific CD4 and CD8 T cells, we also characterized cytokine profiles of IFNγ, IL-2 and TNFα on a single cell level. Based on the gating strategy shown in figure S4, this allowed distinction of seven subpopulations including multifunctional T cells producing all three cytokines. When analyzed across all three groups, the majority of spike-specific CD4 T cells (42.5%) were multifunctional. In contrast, the dominant population among CD8 T cells consisted of dual positive cells producing IFNγ and TNFα (47.1%) followed by 30.6% IFNγ-single positive cells (figure 1F). When the distribution of cytokine producing cells was separately analyzed for each group, significant differences were found between the three groups, which held true for both spike-specific CD4 and CD8 T cells (figure S5). Given the particular ability of the vector-vaccine to induce T cells during priming^7^, it is notable that the two vector-primed groups had the highest percentage of polyfunctional CD4 T cells irrespective of the boosting vaccine. This also held true for the dominant fraction of CD8 T cells co-producing IFNγ and TNFα. In contrast, SEB-reactive cytokine profiles among CD4 and CD8 T cells were similar in all vaccine groups.

Finally, local and systemic adverse events within the first week after primary and secondary vaccination were recorded using a questionnaire (figure 2A). Local reactions such as pain at the injection site and swelling were similar after priming with the vector and mRNA vaccines (figure 2B). In contrast, the vector-vaccine induced significantly more systemic events including fever, chills, gastrointestinal events, headache, fatigue, myalgia or arthralgia. Participants also reported more frequent use of antipyretic drugs (figure 2C). When comparing reactogenicity after secondary vaccination, both local and systemic events were markedly less frequent after homologous vector boosting. In contrast, boosting with an mRNA vaccine was less well tolerated, and the spectrum of local and systemic adverse events was very similar for both vector- and mRNA-primed individuals. Consequently, individual perception on whether vaccine recipients were more severely affected after the first or the second dose was clearly determined by severity of the priming vaccine (figure 2D). Hence, both recipients of the homologous vector regimen and of the heterologous vector/mRNA regimen were most affected by the priming vector vaccination.

**Figure 2:**
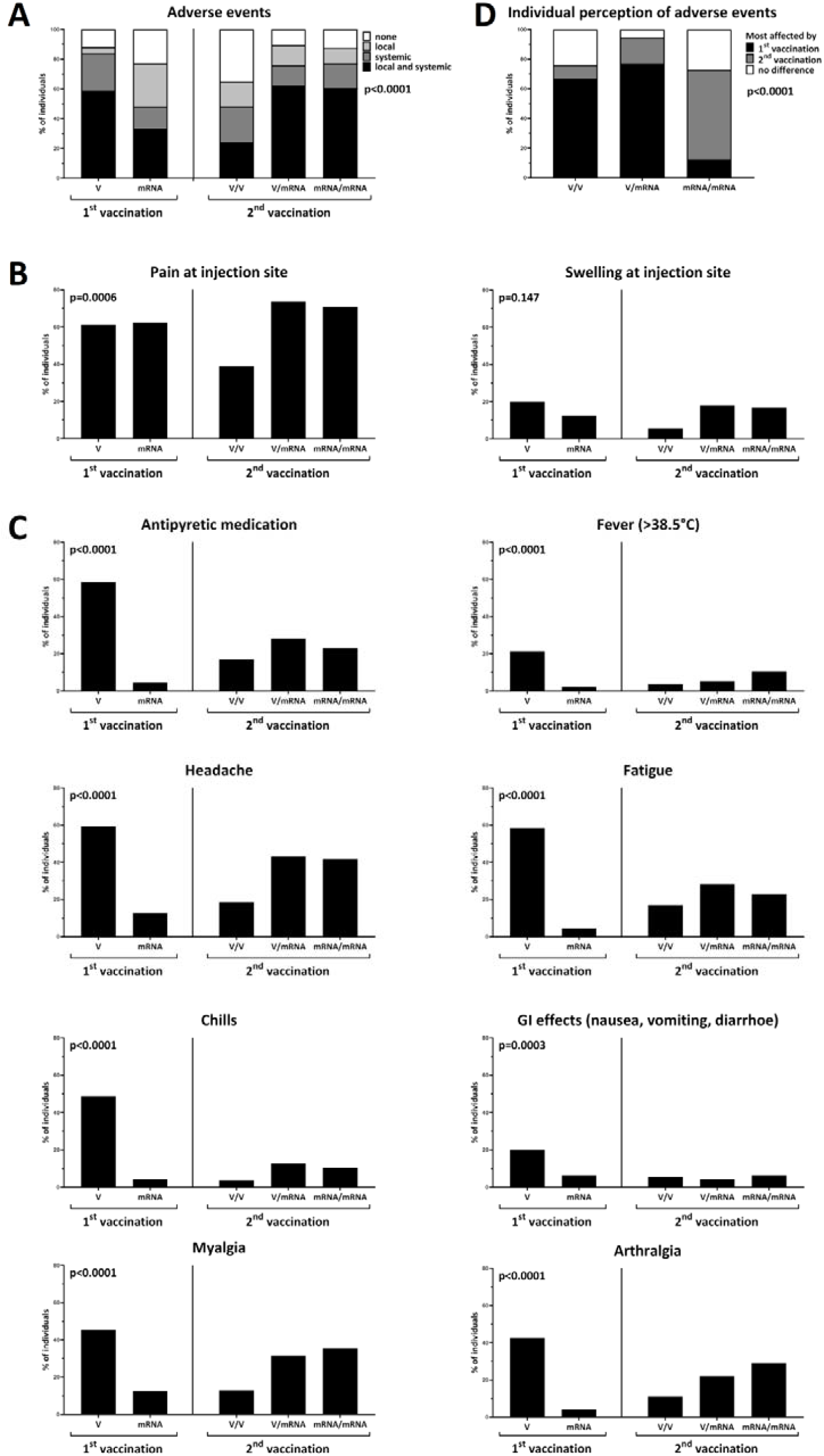
Reactogenicity after primary and secondary vaccination with homologous and heterologous prime-boost regimens. Reactogenicity within the first week after the priming and after the boosting dose was self-reported based on a standardized questionnaire and analyzed after first vaccination with vector (V, n=150) or mRNA vaccine (mRNA, n=48) and after second vaccination with homologous (V/V, n=54; mRNA/mRNA, n=48) and heterologous (V/mRNA, n=95) vaccine regimens with respect to **(A)** local/systemic reactions in general, and stratified for local **(B)** and various systemic adverse events **(C)**. Individual perception of which of the two vaccinations affected more is shown in panel **(D)**. Comparisons between the groups were performed using X^2^ test.

Mixing different vaccine types in heterologous regimens has already been deployed in previous vaccine studies. Examples include experimental vaccines towards HIV^9^ or malaria^10^, and the licensed vaccine against Ebola virus disease, that is based on priming with an adenovirus vector followed by boosting with a modified vaccinia virus vector^11^. Although no data were available on immunogenicity and efficacy of heterologous strategies among licensed COVID-19 vaccines, this raised confidence in recommending a heterologous mRNA booster vaccination in ChAdOx1-vector primed individuals after recognition of severe adverse events of cerebral venous thrombosis^4,5^. We here show that the heterologous regimen led to a strong induction of both antibodies and T cells. IgG-levels were similar in magnitude as after homologous mRNA vaccination and approximately 10-fold higher than IgG-levels after homologous vector vaccination. Similar differences were found for vaccine-induced CD4 T cells, while neutralizing antibody activity and spike-specific CD8 T cells were even more pronounced after heterologous vaccination. Similar results were recently reported in mice^12^. Despite the strong ability of the ChAdOx1 vaccine to induce T cells after priming^7^, the strikingly lower immunogenicity after the homologous ChAdOx1 booster dose affected both antibodies and T cells. This may result from immunity towards the vector induced after the first vaccine dose^13^, which may have rendered secondary vaccination less efficient. Nevertheless, our results show that both vector-primed antibodies and T cells are particularly well induced when combined with mRNA as secondary vaccine. This indicates that the homologous ChAdOx1-regimen is unable to exploit the full potential of the vaccine. However, this vaccine clearly holds promise as a valuable component of a mix-and-match strategy to boost T-cell immunity.

Although the heterologous group suffered from more pronounced systemic adverse events after vector priming, secondary vaccination with the mRNA was less severe and well tolerated, and the spectrum of both local and systemic adverse events was comparable to the homologous mRNA regimens. This contrasts with reactogenicity data from the Com-COV trial, where adverse events in the heterologous regimens were more severe than in homologous groups^14^. Whether this was due to the shorter interval between priming and boosting (4 weeks versus 9-12 weeks in our study) and whether this affects immunogenicity awaits further study.

Our study is limited by the fact that we did not include comparative data on immunity after the first vaccine dose. However, this was addressed in a previous study showing that antibody levels were higher after mRNA-priming whereas T-cell levels were higher after vector-priming^7^. Moreover, despite similar results for BNT162b2 and mRNA-1273, our data are mainly based on the BNT162b2 mRNA vaccine. Finally, our study participants were recruited in a real-word setting, where the homologous ChAdOx1 nCoV-19 vaccine group was slightly older. However, as a subgroup analysis of age matched individuals gave the same results (data not shown), the difference in age unlikely accounts for the strikingly less pronounced immune responses in this group. Finally, no efficacy data of heterologous regimens are available to inform protection from infection or disease. While this awaits further study, definition of immune-based correlates of protection will be important to estimate efficacy of novel vaccines and vaccine combinations^15^. Neutralizing antibodies have been discussed as promising candidates^15,16^ that mirror efficacy of vector and mRNA regimens^1-3^. As immunogenicity after heterologous vaccination is comparable or in part superior to the homologous mRNA-regimens, it will be interesting to see whether this translates into similar efficacy.

Although vaccine development focused on antibodies due to their ability to confer sterilizing immunity, T cells are important in mediating protection from severe disease^17^, and T cells may be less affected by virus variants^18^. In this respect, knowledge on immunity and reactogenicity after heterologous vaccination from this and similar studies may speed immunization campaigns and will have impact on future development of vaccine strategies including how to improve vaccine-induced T-cell immunity and protection from severe disease among vulnerable groups of immunocompromised patients.

## Methods

### Study design and subjects

Immunocompetent individuals with no known history of SARS-CoV-2 infection were included in the study. Individuals were enrolled after having received a homologous vaccine regimen comprising of either ChAdOx1 nCoV-19 or one of the mRNA-vaccines (BNT162b2 or mRNA-1273), or a heterologous vaccine regimen comprising a ChAdOx1 nCoV-19 priming dose followed by secondary vaccination with an mRNA vaccine. The time interval between the first and the second dose was determined as per national guideline and varied from 3-6 weeks for the homologous mRNA regimens to 9-12 weeks for the homologous ChAdOx1 nCoV-19, and the heterologous ChAdOx1 nCoV-19/mRNA regimen^19^. Lymphocyte subpopulations as well as vaccine-induced SARS-CoV-2-specific humoral and cellular immunity were determined from heparinized whole blood 14 days after the second vaccine dose with an interval of 13-18 days tolerated. Local and systemic adverse events within 7 days after the first and the second vaccination were self-reported using a standardized questionnaire. The study was approved by the ethics committee of the Ärztekammer des Saarlandes (reference 76/20), and all individuals gave written informed consent.

### Quantification of lymphocyte populations and plasmablasts

T cells, B cells and plasmablasts were quantified from 100 µl heparinized whole blood as described before^8^ using monoclonal antibodies towards CD3 (clone SK7), CD19 (clone HIB19), CD27 (clone L128), CD38 (clone HB7) and IgD (clone IA6-2). T and B cells were identified among total lymphocytes by expression of CD3 and CD19, respectively. Plasmablasts were characterized by expression of CD38 among IgD-CD27+ CD19 positive switched-memory B cells. CD4 and CD8 T cells were quantified after additional staining of CD4 (clone SK3) and CD8 (clone RPA-T8). Antibodies are listed in table S2. The gating strategy is shown in figure S1. Absolute lymphocyte numbers were calculated based on differential blood counts.

### Quantification of vaccine induced SARS-CoV-2-specific T cells

SARS-CoV-2-specific T cells were determined from heparinized whole blood after a 6h-stimulation with overlapping peptides spanning the SARS-CoV-2 spike protein (N-terminal receptor binding domain and C-terminal portion including the transmembrane domain, each peptide 2µg/ml; JPT, Berlin, Germany) as described previously^8^. Stimulations with 0.64% DMSO and with 2.5μg/ml of *Staphylococcus aureus* enterotoxin B (SEB; Sigma) served as negative and positive controls, respectively. All stimulations were carried out in presence of co-stimulatory antibodies against CD28 and CD49d (1μg/ml each). Immunostaining was performed using anti-CD4 (clone SK3), anti-CD8 (clone SK1), anti-CD69 (clone L78), anti-IFNγ (clone 4S.B3), anti-IL-2 (clone MQ1-17H12), and anti-TNFα (clone MAb11) and analyzed using flow-cytometry. Antibodies are listed in table S2. SARS-CoV-2-reactive CD4 or CD8 T cells were identified as activated CD69 positive T cells producing IFNγ (see figure S2A for gating strategy). Moreover, co-expression of IL-2 and TNFα was analyzed to characterize cytokine expression profiles. Reactive CD4 and CD8 T-cell levels after control stimulations were subtracted from levels obtained after SARS-CoV-2-specific stimulation, and 0.03% of reactive T cells was set as detection limit based on the distribution of T-cell frequencies after control stimulations.

### Determination of SARS-CoV-2-specific antibodies and neutralization capacity

SARS-CoV-2-specific IgG antibodies towards the receptor binding domain of SARS-CoV-2 spike protein were quantified using an enzyme-linked immunosorbent assay (ELISA) according to the manufacturer’s instructions (SARS-CoV-2-QuantiVac, Euroimmun, Lübeck, Germany). Antibody binding units (BAU/ml) <25.6 were scored negative, ≥25.6 and <35.2 were scored intermediate, and ≥35.2 were scored positive. A neutralization assay based on antibody-mediated inhibition of soluble ACE2 binding to the plate-bound S1 receptor binding domain was used according to the manufacturer’s instructions (SARS-CoV-2-NeutraLISA, Euroimmun, Lübeck, Germany). Neutralizing capacity was calculated as percentage of inhibition (IH) by 1 minus the ratio of the extinction of the respective sample and the extinction of the blank value. IH<20% was scored negative, IH≥20 to <35 intermediate, and IH≥35% positive.

### Statistical analysis

Kruskall Wallis test, followed by Dunn’s multiple comparison test, was performed to compare unpaired non-parametric data between groups (lymphocyte subpopulations, T-cell and antibody levels). Data with normal distribution were analyzed using an unpaired ANOVA (cytokine expression profiles, age). Categorial analyses on gender and adverse events were performed using X^2^ test. Correlations between levels of T cells, antibodies, and plasmablasts were analyzed according to Spearman. A p-value <0.05 was considered statistically significant. Analysis was carried out using GraphPad Prism 9.0 software (GraphPad, San Diego, CA, USA) using two-tailed tests. Cytokine profiles were plotted using the VennDiagram package (version 1.6.20)^20^ running under R (version 4.0.2).

## Data Availability

All figures have associated raw data. The data that support the findings of this study are available from the corresponding author upon reasonable request.

## Author Contributions

T.S., D.S., U.S., B.C.G., S.S., and M.S. designed the study; T.S., D.S., U.S. and M.S. designed the experiments, V.K., F.H., S.M., A.A.-O., and T.S. performed experiments; S.S., B.C.G., J.M., S.L.B. and U.S. contributed to study design, patient recruitment, and clinical data acquisition. D.S., V.K., T.S., U.S., J.M. and M.S. supervised all parts of the study, performed analyses and wrote the manuscript. All authors approved the final version of the manuscript.

## Acknowledgements

The authors thank Candida Guckelmus and Rebecca Urschel for excellent technical assistance, and Dr. Christina Baum and the team of the occupational health care center at Saarland University medical center, and Susanne Brehmer and Inna Vallar for their valuable support in enrolling participants. The authors also thank all participants to this study who contributed to the gain in knowledge from this project. Financial support was given by the State Chancellery of the Saarland.

## Abbreviations

BAU: antibody binding units
COVID-19: coronavirus disease 2019
DL: detection limit
ELISA: enzyme-linked immunosorbent assay
IFN: interferon
IH: percentage of inhibition
IL: interleukin
SARS-CoV-2: Severe acute respiratory syndrome coronavirus type 2
SEB: *Staphylococcus aureus* enterotoxin B
TNF: tumor necrosis factor

## References

1. Voysey, M., et al. Safety and efficacy of the ChAdOx1 nCoV-19 vaccine (AZD1222) against SARS-CoV-2: an interim analysis of four randomised controlled trials in Brazil, South Africa, and the UK. Lancet 397, 99–111 (2021).

2. Baden, L.R., et al. Efficacy and Safety of the mRNA-1273 SARS-CoV-2 Vaccine. N Engl J Med 384, 403–416 (2021).

3. Polack, F.P., et al. Safety and Efficacy of the BNT162b2 mRNA Covid-19 Vaccine. N Engl J Med 383, 2603–2615 (2020).

4. Greinacher, A., et al. Thrombotic Thrombocytopenia after ChAdOx1 nCov-19 Vaccination. N Engl J Med 384, 2092–2101 (2021).

5. Schultz, N.H., et al. Thrombosis and Thrombocytopenia after ChAdOx1 nCoV-19 Vaccination. N Engl J Med 384, 2124–2130 (2021).

6. Vygen-Bonnet, S., et al. Beschluss der STIKO zur 5. Aktualisierung der COVID-19-Impfempfehlung und die dazugehörige wissenschaftliche Begründung. Epid Bull 19, 24–36 (2021).

7. Schmidt, T., et al. Cellular immunity predominates over humoral immunity after the first dose of COVID-19 vaccines in solid organ transplant recipients. medRxiv (2021).

8. Schub, D., et al. High levels of SARS-CoV-2-specific T cells with restricted functionality in severe courses of COVID-19. JCI Insight 5(2020).

9. Rerks-Ngarm, S., et al. Vaccination with ALVAC and AIDSVAX to prevent HIV-1 infection in Thailand. N Engl J Med 361, 2209–2220 (2009).

10. McConkey, S.J., et al. Enhanced T-cell immunogenicity of plasmid DNA vaccines boosted by recombinant modified vaccinia virus Ankara in humans. Nat Med 9, 729–735 (2003).

11. Pollard, A.J., et al. Safety and immunogenicity of a two-dose heterologous Ad26.ZEBOV and MVA-BN-Filo Ebola vaccine regimen in adults in Europe (EBOVAC2): a randomised, observer-blind, participant-blind, placebo-controlled, phase 2 trial. Lancet Infect Dis 21, 493–506 (2021).

12. Spencer, A.J., et al. Heterologous vaccination regimens with self-amplifying RNA and adenoviral COVID vaccines induce robust immune responses in mice. Nat Commun 12, 2893 (2021).

13. Ramasamy, M.N., et al. Safety and immunogenicity of ChAdOx1 nCoV-19 vaccine administered in a prime-boost regimen in young and old adults (COV002): a single-blind, randomised, controlled, phase 2/3 trial. Lancet 396, 1979–1993 (2021).

14. Shaw, R.H., et al. Heterologous prime-boost COVID-19 vaccination: initial reactogenicity data. Lancet 397, 2043–2046 (2021).

15. Kristiansen, P.A., et al. WHO International Standard for anti-SARS-CoV-2 immunoglobulin. Lancet 397, 1347–1348 (2021).

16. Khoury, D.S., et al. Neutralizing antibody levels are highly predictive of immune protection from symptomatic SARS-CoV-2 infection. Nat Med (2021).

17. Rydyznski Moderbacher, C., et al. Antigen-Specific Adaptive Immunity to SARS-CoV-2 in Acute COVID-19 and Associations with Age and Disease Severity. Cell 183, 996–1012 e1019 (2020).

18. Skelly, D.T., et al. Vaccine-induced immunity provides more robust heterotypic immunity than natural infection to emerging SARS-CoV-2 variants of concern. Research Square (2021).

19. Vygen-Bonnet, S., et al. Beschluss der STIKO zur 3. Aktualisierung der COVID-19-Impfempfehlung und die dazugehörige wissenschaftliche Begründung. Epid Bull 12, 13–25 (2021).

20. Chen, H. & Boutros, P.C. VennDiagram: a package for the generation of highly-customizable Venn and Euler diagrams in R. BMC Bioinformatics 12, 35 (2011).

