## Supplementary information (tables S1/S2 and figures S1-S5) for "Immunogenicity and reactogenicity of a heterologous COVID-19 prime-boost vaccination compared with homologous vaccine regimens"

### Supplementary tables

#### Table S1: Demographic and clinical characteristics of the study population

| 1° vaccine  2° vaccine | **Vector**  **vector^1^** | **Vector**  **mRNA^2^** | **mRNA**  **mRNA^3^** |  |
| --- | --- | --- | --- | --- |
|  | n=55 | n=97 | n=64 | p-value |
| **Years of age (mean±SD)** | 48.6±11.9 | 40.7±11.1 | 44.5±14.1 | 0.001 |
| **Female gender, n (%)** | 35 (63.6) | 71 (73.2) | 44 (68.8) | 0.465 |
| **Weeks between 1° and 2° vaccination, (mean±SD)** | 10.8±1.4 | 11.2±1.3 | 4.3±1.1 |  |
| **Analysis time [days after 2° vaccination],**  **median (IQR)** | 14 (2) | 14 (1) | 14 (1.75) |  |
| **Differential blood cell counts** | n=55 | n=97 | n=62 |  |
| Leukocytes (cells/µl), median (IQR) | 6800 (2500) | 6900 (2200) | 6950 (2625) | 0.927 |
| Granulocytes (cells/µl), median (IQR) | 4057 (1829) | 3861 (1829) | 4062 (1776) | 0.878 |
| Monocytes (cells/µl), median (IQR) | 590 (238) | 545 (258) | 558 (177) | 0.228 |
| Lymphocytes (cells/µl), median (IQR) | 2291 (840) | 2230 (708) | 2214 (822) | 0.942 |
| CD3 T cells (cells/µl),  median (IQR)^#^ | 1565 (603) | 1594 (635) | 1559 (878) | 0.914 |
| CD4 T cells (cells/µl),  median (IQR) ^#^ | 1000 (450) | 974 (387) | 1065 (615) | 0.398 |
| CD8 T cells (cells/µl),  median (IQR) ^#^ | 394 (282) | 432 (242) | 378 (274) | 0.270 |
| CD19 B cells (cells/µl),  median (IQR)^#^ | 218 (193) | 200 (140) | 200 (142) | 0.750 |

^1^55x ChAdOx1 nCoV-19; ^2^96x BNT162b2, 1x mRNA-1273; ^3^54x BNT162b2, 10x mRNA-1273; ^#^B and T-cell counts were calculated on 55 vector-vector, 97 vector-mRNA and 34 mRNA-mRNA vaccinated individuals, CD4 and CD8 T-cell counts in subgroups of 55, 97 and 16 individuals, respectively.

#### Table S2: Antibodies for flow-cytometric analyses

| **Antigen** | **conjugate** | **clone** | **isotype** | **reactivity** |
| --- | --- | --- | --- | --- |
| CD3 | PerCP | SK7 | IgG1 k | mouse anti-human |
| CD4 | APC-H7 | SK3 | IgG1 k | mouse anti-human |
| CD8 | V500, PerCP | RPA-T8, SK1 | IgG1 k | mouse anti-human |
| CD19 | FITC | HIB19 | IgG1 k | mouse anti-human |
| CD27 | APC | L128 | IgG1 k | mouse anti-human |
| CD38 | PE | HB7 | IgG1 k | mouse anti-human |
| CD69 | PE-Cy7 | L78 | IgG1 k | mouse anti-human |
| IFNγ | FITC | 4S.B3 | IgG1 k | mouse anti-human |
| IgD | PE-Cy7 | IA6-2 | IgG2a k | mouse anti-human |
| IL-2 | PE | MQ1-17H12 | IgG2a k | rat anti-human |
| TNFα | V450 | MAb11 | IgG1 k | mouse anti-human |

All antibodies were purchased from BD.

### Supplementary figures

#### Figure S1: Gating strategy for analysis of T cells and plasmablasts. CD3 positive T cells were identified among single lymphocytes and further subclassified in CD4 and CD8 T cells. Among single lymphocytes, B cells were identified by CD19 expression and subclassified in naïve, non-switched memory and switched memory B cells according to expression of IgD and CD27. Plasmablasts were identified by high expression of CD38 among switched memory B cells.

#### Figure S2: Representative analysis of spike- and SEB-reactive CD4 and CD8 T cells. (A) Lymphocytes were identified among total events by backgating of CD4 and/or CD8 positive cells combined with signals for size (FSC) and granularity (SSC). Hight and area signals of FSC were used to exclude doublets. CD4 T cells were identified among single cells by CD4 positive and CD8 negative signals. Likewise, CD8 T cells were defined as T cells being CD8 positive and CD4 negative. In (B) and (C) representative contour plots of CD4 and CD8 T cells of a 37-years old female are shown after antigen-specific stimulation with SARS-CoV-2 spike peptides or respective control stimuli for negative (DMSO) or positive control (SEB) stimulation. Numbers indicate percentages of CD4 or CD8 T cells co-expressing the activation marker CD69 and the cytokines IFNγ, IL-2 and/or TNFα. FSC, forward scatter; IFN, interferon; IL, interleukin; SEB, *Staphylococcus aureus* enterotoxin B; SSC, side scatter; TNF, tumor necrosis factor.

#### Figure S3: SARS-CoV-2 spike-specific T-cell responses producing TNFα and IL-2 after vaccination with homologous and heterologous prime-boost regimens. Percentages of SARS-CoV-2 spike-specific CD4 and CD8 T cells were determined after antigen-specific stimulation of whole blood samples followed by intracellular cytokine analysis using flow-cytometry. Reactive cells were identified by co-expression of the activation marker CD69 and the cytokine tumor necrosis factor (TNF) α (left panel), interleukin 2 (IL-2, middle panel), or any of the cytokines analyzed (IFNγ / TNFα / IL-2, right panel), respectively, among CD4 or CD8 T cells and subtraction of background reactivity of respective negative control stimulations. T-cell responses were compared between individuals who either received SARS-CoV-2 vector/vector (V/V, n=55), vector/mRNA (V/mRNA, n=97) or mRNA/mRNA vaccines (mRNA/mRNA, n=64) using Kruskal-Wallis test with Dunn’s multiple comparisons post test. Bars represent medians with interquartile ranges, individuals who received the mRNA-1273 vaccine are indicated by grey symbols. *p<0.05, ****p<0.0001.

#### Figure S4: Gating strategy for analysis of cytokine expression profiles. To characterize spike- and SEB-reactive CD4 T cells regarding their single or combined expression of the cytokines IFNγ, IL-2, and TNFα, CD4 T cells positive for combined expression of CD69 and IFNγ were divided into four subpopulations according to additional expression of IL-2 and TNFα. Using NOT Boolean Gating, all CD4 T cells which were not CD69+IFNγ+ were analyzed for CD69+IL-2+ and CD69+TNFα+ CD4 T cells. Using OR Boolean Gating, CD69+IL-2+ and/or CD69+TNFα+ CD4 T cells were divided into IL-2 single, TNFα single or IL-2+TNFα+ cells. After subtraction of background reactivity from negative control stimulations, the sum of these 7 subpopulations was set to 100%. A similar strategy was applied for CD8 T cells. IFN, interferon; IL, interleukin; TNF, tumor necrosis factor.

#### Figure S5: Cytokine expression profiles of antigen-specific CD4 and CD8 T cells after vaccination with homologous and heterologous prime-boost regimens. Cytokine expression of CD4 (A) and CD8 T cells (B) after stimulation with overlapping peptides of SARS-CoV-2 spike protein (upper panels) or *Staphylococcus aureus* enterotoxin B (SEB), lower panels) was compared between individuals who either received SARS-CoV-2 vector/vector (V/V), vector/mRNA (V/mRNA), or mRNA/mRNA vaccine combinations (mRNA/mRNA, n=64). Cytokine expressing T cells were divided into 7 subpopulations according to their expression of IFNγ, IL-2, and TNFα (single, double or triple cytokine expressing cells, for gating strategy, see figure S4). The samples from all study participants were analyzed. To ensure robust statistical analysis in this setting, analysis was restricted to samples with at least 30 cytokine expressing CD4 or CD8 T cells, respectively (n=31 and 15 for V/V, n=90 and 73 for V/mRNA and n=60 and 25 for mRNA/mRNA vaccine regimes). Differences among subpopulations between the groups were determined using ordinary one-way ANOVA test. Bars represent means and standard deviations of subpopulations among all SARS-CoV-2 spike-specific or SEB-reactive CD4 T cells. IFN, interferon, IL, interleukin, TNF, tumor necrosis factor; *p<0.05, **p<0.01, ***p<0.001, ****p<0.0001.

### Supplementary figures

#### Supplementary figure S1

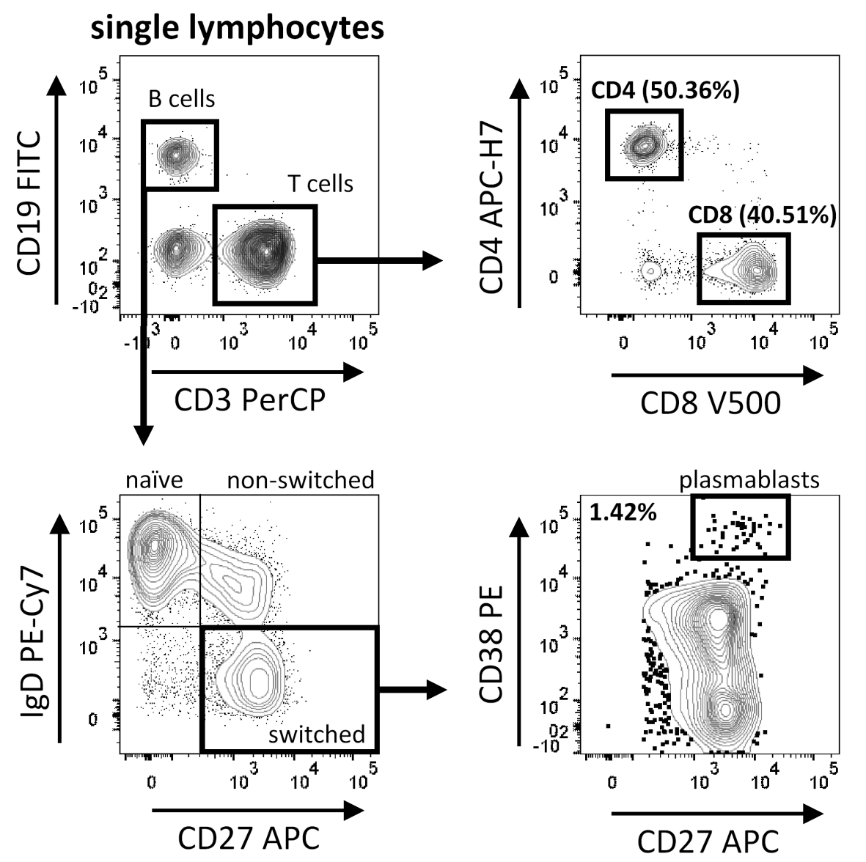

#### Supplementary figure S2

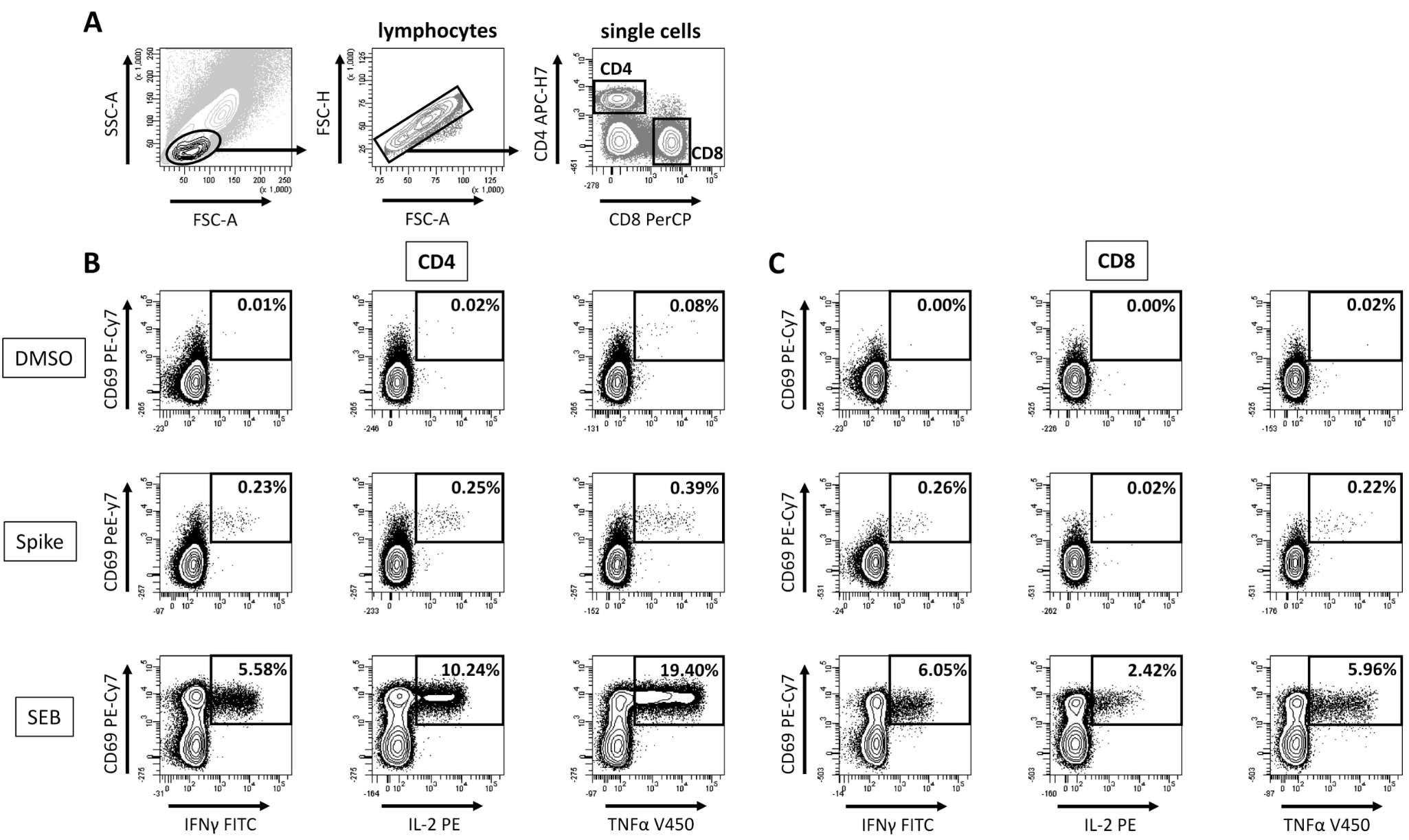

#### Supplementary figure S3

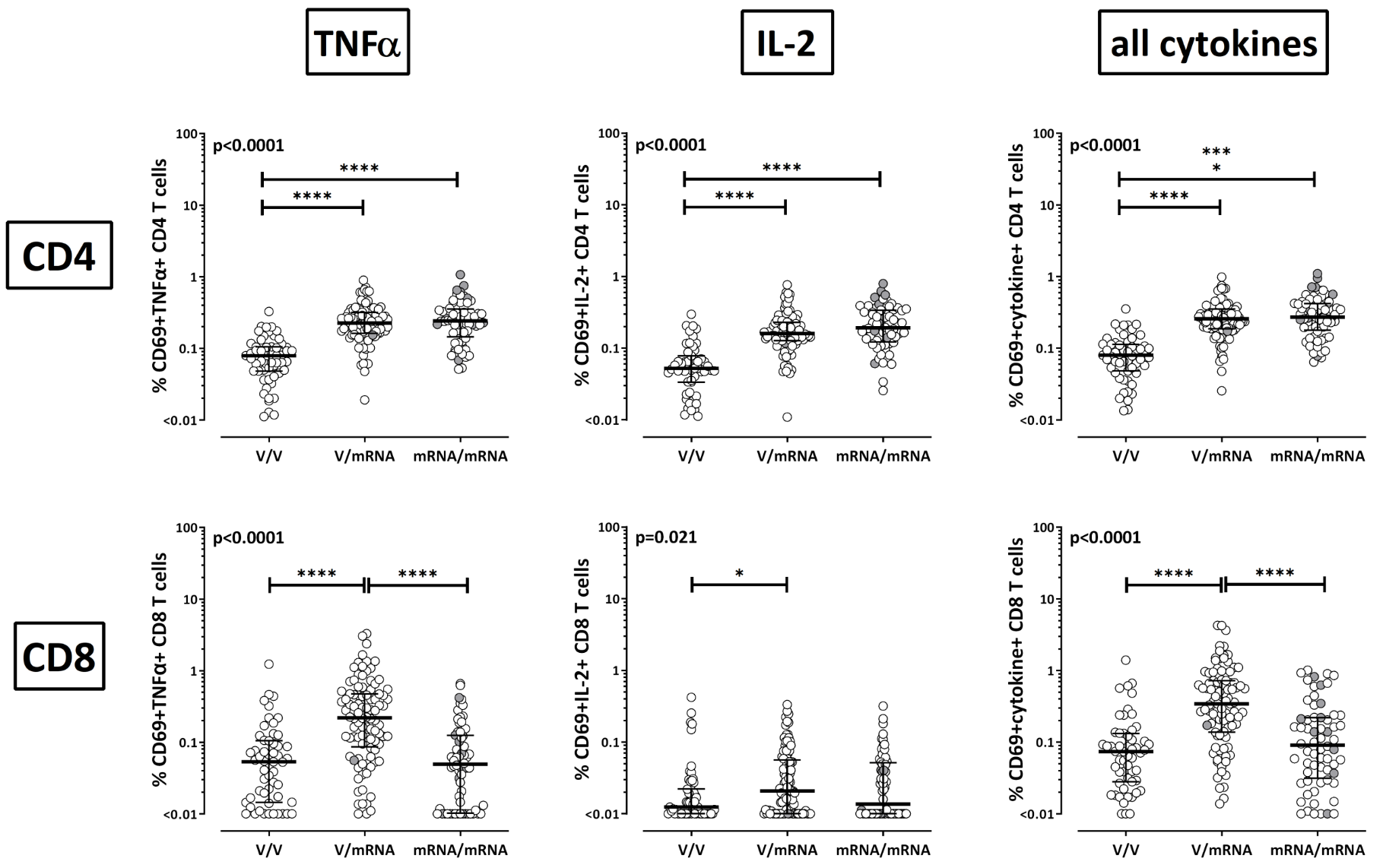

#### Supplementary figure S4

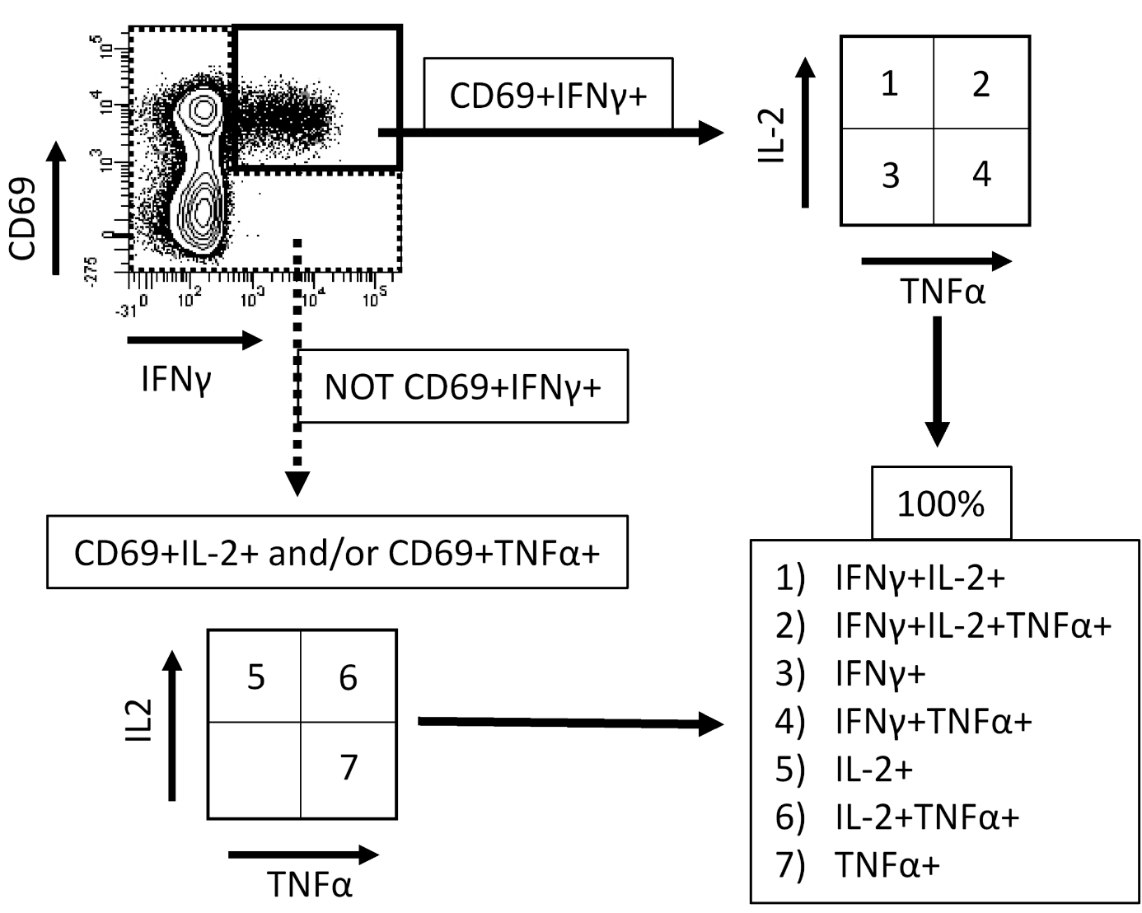

#### Supplementary figure S5

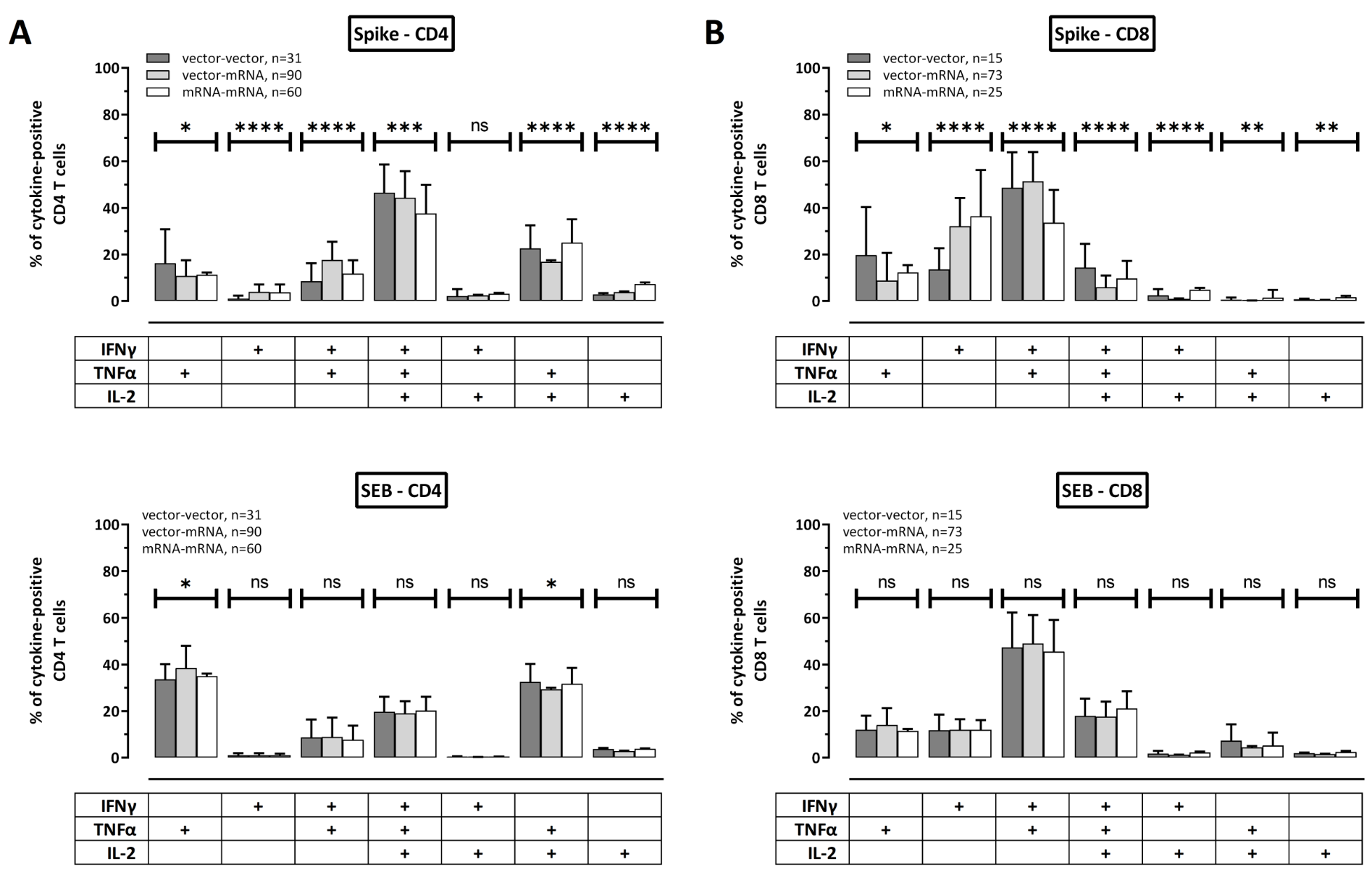
